# The ‘fat but powerful’ paradox: association of muscle power and adiposity markers with all-cause mortality in older adults from the EXERNET Multi-center Study

**DOI:** 10.1101/2020.11.13.20231092

**Authors:** Julian Alcazar, David Navarrete-Villanueva, Asier Mañas, Alba Gómez-Cabello, Raquel Pedrero-Chamizo, Luis M. Alegre, Jose G. Villa-Vicente, Narcis Gusi, Marcela González-Gross, Jose A. Casajús, German Vicente-Rodríguez, Ignacio Ara

## Abstract

**Objectives:** To assess the influence of muscle power and adiposity on all-cause mortality risk and to evaluate the ‘fat but powerful’ (or ‘fat but fit’) paradox in older adults.

**Methods:** A total of 2563 older adults (65‒91 years old) from the EXERNET Multi-center study were included. Adiposity (body mass index (BMI), waist circumference, body fat percentage (BF%) and fat index), allometric and relative power (sit-to-stand muscle power test) and various covariates (age, sex, hypertension, smoking status, and walking and sitting times per day) were registered at baseline. All-cause mortality was recorded during a median follow-up of 8.9 years. Participants were classified into four groups: lean and powerful (L+P), fat but powerful (F+P), lean but weak (L+W) and fat and weak (F+W). Cox proportional hazard regression models and adjusted hazard ratios (HR) were calculated.

**Results:** According to BMI and waist circumference, all-cause mortality risk was reduced in the F+P (HR=0.55 and 0.63, respectively; p≤0.049) and L+P (HR=0.57 and 0.58, respectively; p≤0.043) groups. According to BF%, all-cause mortality decreased in the L+P group (HR=0.53; p=0.021), and a trend for a reduction was reported in the F+P group (HR=0.57; p=0.060). According to fat index, a survival benefit was only noted in the L+P group (HR=0.50; p=0.049). Higher levels of relative power reduced all-cause mortality risk among older people (HR=0.63 and 0.53, respectively; p≤0.011).

**Conclusion:** Powerful older people exhibited a reduced 9-year all-cause mortality regardless of BMI, waist circumference and BF%. Obesity according to fat index blunted the survival benefits of being powerful.

## 1. Introduction

Physical fitness has been identified as one of the most important predictors of all-cause mortality among adults. Specifically, low cardiorespiratory fitness has been shown to be responsible for a greater number of deaths than other risk factors such as hypertension, smoking or metabolic disease ^1,2^. The relevance of achieving an adequate physical fitness status is such that people with obesity or metabolic disease who present adequate cardiorespiratory fitness exhibit a diminished mortality risk when compared to people without obesity or metabolic disease but presenting low cardiorespiratory fitness ^3^. This has been described as the ‘fat but fit’ paradox ^4^.

The ‘fat but fit’ paradox has also been confirmed in older adults. Sui et al. ^5^ demonstrated that maintaining an adequate cardiorespiratory fitness at older age decreases all-cause mortality risk independently of overall or abdominal adiposity. However, the assessment of cardiorespiratory fitness demands considerable time and resources from the practitioners and requires an effort until exhaustion from the older participants. Among the other components of physical fitness, muscle power (i.e. product of force and velocity) has been found to be more strongly related with older adults’ functional status than cardiorespiratory capacity ^6^. Muscle power has also been reported to predict all-cause mortality among men, independently of physical activity, muscle mass and strength ^7^. Fortunately, muscle power can be assessed using a feasible procedure that only requires a chair and a stopwatch and a few seconds to be conducted: the sit-to-stand (STS) muscle power test ^8,9^. Nevertheless, no studies have previously assessed the ‘fat but fit’ paradox while considering muscle power as the physical fitness component, which might be of special relevance in older populations. Hence, the main goals of the present investigation were twofold: i) to evaluate and compare all-cause mortality risk in a representative sample of non-institutionalized Spanish older adults presenting different combinations of adiposity and muscle power; and ii) to determine whether the ‘fat but fit’ paradox occurs when muscle power is regarded.

## 2. Methods

### 2.1 Study design

This is a prospective study conducted in the participants of the EXERNET multi-center study ^10,11^. This study includes a representative sample of non-institutionalized older adults (≥65 years old) living in Spain. Data was collected from June 2008 to November 2009 by means of personal interviews and a physical examination (body composition and physical fitness). All the participants gave their written informed consent. The study was approved by the Clinical Research Ethics Committee of Aragón (18/2008) and the Ethical Committee of the University Hospital Fundación Alcorcón (50/2016), and the procedures were performed in accordance with the Helsinki Declaration.

### 2.2 Participants

A total of 2563 older adults (602 older men and 1961 older women) were included in this study. Inclusion criteria were age ≥65 years old and independent living. Older people with dementia, cancer or terminal illness were excluded. Baseline information of the study participants is shown in Table 1.

**Table 1.** Baseline characteristics of the study participants.

|  | Men (n=602) |  |  | Women (n=1961) |  |  |
| --- | --- | --- | --- | --- | --- | --- |
|  | Mean | ± | SD | Mean | ± | SD |
| Age (years) | 72.1 | ± | 5.4 | 72.0 | ± | 5.2 |
| Body mass (kg) | 77.6 | ± | 11.0 | 68.9 | ± | 10.7 |
| Height (m) | 1.65 | ± | 6.51 | 1.53 | ± | 5.8 |
| BMI (kg·m<sup>-2</sup>) | 28.3 | ± | 3.5 | 29.5 | ± | 4.3 |
| Waist circumference (cm) | 99.2 | ± | 10.0 | 93.0 | ± | 12.3 |
| Body fat (%) | 29.1 | ± | 5.1 | 39.7 | ± | 5.2 |
| Fat index (kg·m<sup>-2</sup>) | 8.3 | ± | 2.2 | 11.9 | ± | 3.1 |
| STS performance (reps) | 15.0 | ± | 3.6 | 14.1 | ± | 3.3 |
| Allometric power (W·m<sup>-2</sup>) | 99.8 | ± | 26.0 | 80.8 | ± | 20.4 |
| Relative power (W·kg<sup>-1</sup>) | 3.54 | ± | 0.83 | 2.76 | ± | 0.67 |
| Hypertension (Yes, %) | 52.6 |  |  | 50.2 |  |  |
| Smoking (Yes, %) | 9.0 |  |  | 1.7 |  |  |
| Walking <1 h·day<sup>-1</sup> (Yes, %) | 28.2 |  |  | 35.5 |  |  |
| Sitting ≥5 h·day<sup>-1</sup> (Yes, %) | 18.3 |  |  | 14.5 |  |  |

*Note:* BMI, body mass index. SD, standard deviation. STS, sit-to-stand.

### 2.3 Anthropometrics and body composition

Subjects remained in the standing position without shoes and shocks while using light clothing. Any metal object was removed. Height and body mass were assessed with a portable stadiometer and scale device (SECA 225, SECA, Germany), and body mass index (BMI; kg·m^-2^) was calculated as the ratio between body mass and height squared. Waist circumference (cm) was measured at the narrowest point between the lower border of the last rib and the iliac crest with an inelastic measuring tape (Rosscraft Innovations Inc., Canada). Whole body fat was estimated using a bioelectrical impedance analyzer (Tanita BC-418 MA, Tanita Corp., Japan). Body fat percentage (%) was calculated as the ratio between body fat and body mass. Fat index (kg·m^-2^) was calculated as the ratio between body fat and height squared. Then, obesity was identified separately in men and women for the different adiposity indexes: BMI ≥30 kg·m^-2^ in both men and women ^12^; waist circumference ≥101 cm in men and ≥88 cm in women ^13^; body fat percentage ≥31% in men and ≥43% in women ^14^; and fat index ≥9.05 kg·m^-2^ in men and ≥13.06 kg·m^-2^ in women (highest sex-specific quintiles).

### 2.4 Muscle power

Muscle power was evaluated with the 30-s sit-to-stand (STS) muscle power test ^9^. This test consists in performing as many STS repetitions as possible within 30 s on a standardized armless chair. Then, muscle power is calculated by means of a validated equation that considers the participant’s body mass and height, chair height (0.43 m) and 30-s STS performance. Strong verbal encouragement was given to all the participants throughout the test. Due to absolute muscle power is positively associated with height (r = 0.48 in men and 0.47 in women), allometric muscle power (W·m^-2^) was calculated (i.e. absolute muscle power normalized to height squared) ^15^. Low allometric power was identified in men with values <75.4 W·m^-2^ and in women with values <61.5 W·m^-2^ (lowest sex-specific quintile), while values equal or above these cut-off points were regarded normal. In addition, relative muscle power (W·kg^-1^) is a measure of muscle power that integrates information on physical fitness and adiposity levels due to can be calculated as allometric muscle power normalized to BMI. Of note, relative muscle power can also be obtained from the ratio between absolute muscle power and body mass. Low relative power was defined as showing values <2.6 W·kg^-1^ in men and <2.1 W·kg^-1^ in women (lowest sex-specific quintile), otherwise relative muscle power was regarded normal. In addition, high relative power was identified in men with values ≥4.0 W·kg^-1^ and in women with values ≥3.2 W·kg^-1^ (highest sex-specific quintile).

### 2.5 Covariates

Resting systolic blood pressure was assessed with the participants in the sitting position and after 10 min of rest. The average of 2 measurements separated by 2 min was obtained for further analysis. Hypertension was considered in participants with systolic blood pressure ≥140 mmHg. Smoking status, walking time and sedentary time were self-reported by the participants during the personal interviews through a validated questionnaire ^16^. Subjects were categorized into two groups of smoking status (yes and no), two groups of physical activity (walking ≥1 h and <1 h per day; based on the highest quintile) and two groups of sedentary time (sitting ≥5 h and <5 h per day; based on the highest quintile).

### 2.6 Survival data

All-cause mortality was collected in the study participants by consulting the Spanish National Death Index during a median follow-up of 8.9 years (interquartile range = 8.5‒9.1 years) that elapses from the date of interview until date of death or censoring on March 31, 2018 (whichever came first).

### 2.7 Statistical analyses

Data were presented as mean ± standard deviation (SD) or 95% confidence intervals (*lower limit* to *higher limit*) for continuous variables and as percentage for categorical variables. Differences between deceased and surviving participants were assessed separately in men and women using paired t-tests for continuous variables and Chi-Square (χ^2^) tests for categorical variables. To assess the ‘fat but powerful’ paradox, four mutually exclusive groups were created for each adiposity variable: 1) lean and powerful (L+P) (non-obese people with normal allometric power); 2) fat but powerful (F+P) (obese people with normal allometric power); 3) lean but weak (L+W) (non-obese people with low allometric power); and 4) fat and weak (F+W) (obese people with low allometric power). Finally, hazard ratios of all-cause mortality for each corresponding group of adiposity and allometric power [reference: F+W group] were calculated using Cox proportional hazards regression models adjusted for age, sex [reference: men], hypertension, smoking status, walking <1 h·day^-1^ and sitting ≥5 h·day^-1^ [all reference: no]). Deaths within the first two years were excluded in order to minimize bias from reverse causation ^17^. In addition, hazard ratios of all-cause mortality for each group of relative muscle power were obtained adjusting for the same covariates. All statistical analyses were performed using SPSS v20 (SPSS Inc., USA) and the level of significance was set at α = 0.05.

## 3. Results

### 3.1 Comparison between deceased and surviving older people

A total of 215 (9.3%) participants died (86 (15.8%) men and 129 (7.3%) women) during the 9-year follow-up. Deceased men were older (mean difference [95% CI]: 3.8 [2.4 to 5.1] years old), presented lower allometric and relative muscle power values at baseline (‒8.8 [‒14.7 to ‒3.0] W·m^-2^ and ‒0.26 [‒0.45 to ‒0.08] W·kg^-1^, respectively), and were less likely to have hypertension (χ^2^ = 8.246; p = 0.004) and more likely to be smokers (χ^2^ = 4.714; p = 0.030) when compared to surviving men (Table 2). No differences were noted between deceased and surviving men regarding BMI, waist circumference, body fat percentage, fat index, STS performance, and walking and sitting time groups (all p>0.05). Among women, those who died were older (4.5 [3.6 to 5.4] years old), exhibited lower values of STS performance (‒1.0 [‒1.7 to ‒0.4] reps), allometric power (‒8.1 (‒11.8 to ‒4.5) W·m^-2^) and relative power (‒0.28 [‒0.41 to ‒0.14) W·kg^-1^), and were more likely to have hypertension (χ^2^ = 4.984; p = 0.026) than the surviving women (Table 2). No differences were found between deceased and surviving women in terms of BMI, waist circumference, body fat percentage, fat index, walking and sitting time groups and smoking status (all p>0.05).

**Table 2.**
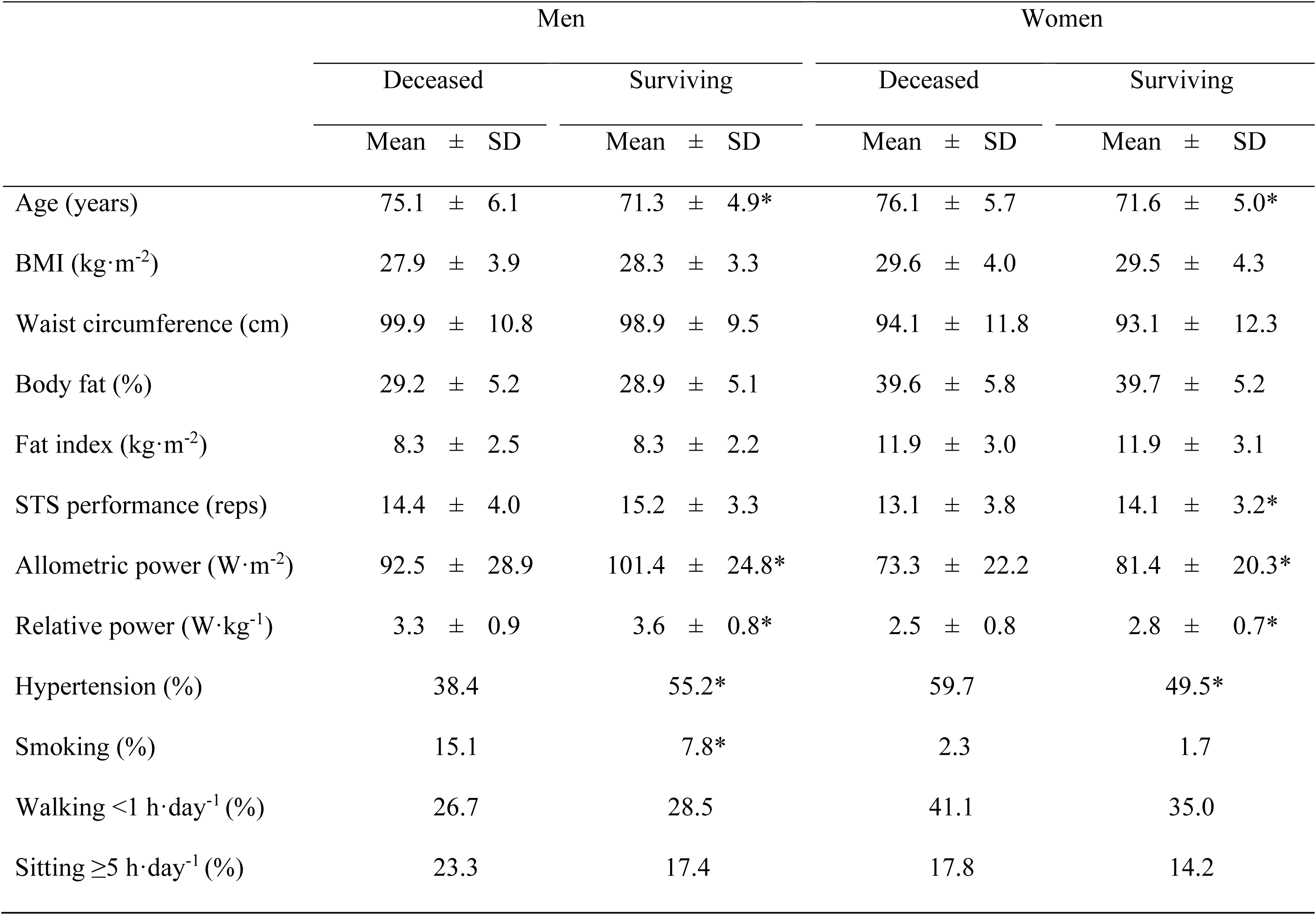
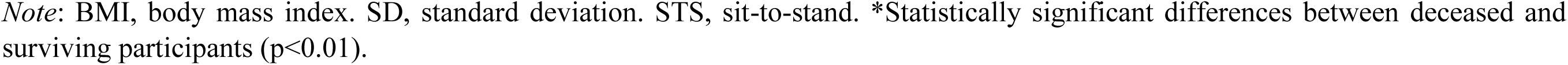
Comparison between deceased and surviving older men and women.

|  | Men |  |  |  |  |  | Women |  |  |  |  |  |
| --- | --- | --- | --- | --- | --- | --- | --- | --- | --- | --- | --- | --- |
|  | Deceased |  |  | Surviving |  |  | Deceased |  |  | Surviving |  |  |
|  | Mean | ± | SD | Mean | ± | SD | Mean | ± | SD | Mean | ± | SD |
| Age (years) | 75.1 | ± | 6.1 | 71.3 | ± | 4.9* | 76.1 | ± | 5.7 | 71.6 | ± | 5.0* |
| BMI (kg·m<sup>-2</sup>) | 27.9 | ± | 3.9 | 28.3 | ± | 3.3 | 29.6 | ± | 4.0 | 29.5 | ± | 4.3 |
| Waist circumference (cm) | 99.9 | ± | 10.8 | 98.9 | ± | 9.5 | 94.1 | ± | 11.8 | 93.1 | ± | 12.3 |
| Body fat (%) | 29.2 | ± | 5.2 | 28.9 | ± | 5.1 | 39.6 | ± | 5.8 | 39.7 | ± | 5.2 |
| Fat index (kg·m<sup>-2</sup>) | 8.3 | ± | 2.5 | 8.3 | ± | 2.2 | 11.9 | ± | 3.0 | 11.9 | ± | 3.1 |
| STS performance (reps) | 14.4 | ± | 4.0 | 15.2 | ± | 3.3 | 13.1 | ± | 3.8 | 14.1 | ± | 3.2* |
| Allometric power (W·m<sup>-2</sup>) | 92.5 | ± | 28.9 | 101.4 | ± | 24.8* | 73.3 | ± | 22.2 | 81.4 | ± | 20.3* |
| Relative power (W·kg<sup>-1</sup>) | 3.3 | ± | 0.9 | 3.6 | ± | 0.8* | 2.5 | ± | 0.8 | 2.8 | ± | 0.7* |
| Hypertension (%) | 38.4 |  |  | 55.2* |  |  | 59.7 |  |  | 49.5* |  |  |
| Smoking (%) | 15.1 |  |  | 7.8* |  |  | 2.3 |  |  | 1.7 |  |  |
| Walking <1 h·day<sup>-1</sup> (%) | 26.7 |  |  | 28.5 |  |  | 41.1 |  |  | 35.0 |  |  |
| Sitting ≥5 h·day<sup>-1</sup> (%) | 23.3 |  |  | 17.4 |  |  | 17.8 |  |  | 14.2 |  |  |

*Note:* BMI, body mass index. SD, standard deviation. STS, sit-to-stand. \*Statistically significant differences between deceased and surviving participants ( $p < 0.01$ ).

### 3.2 Comparisons among groups of power and adiposity regarding all-cause mortality

Survival plots showing the cumulative probability of survival among the different groups of allometric muscle power and adiposity are displayed in Figure 1.

**Figure 1.**
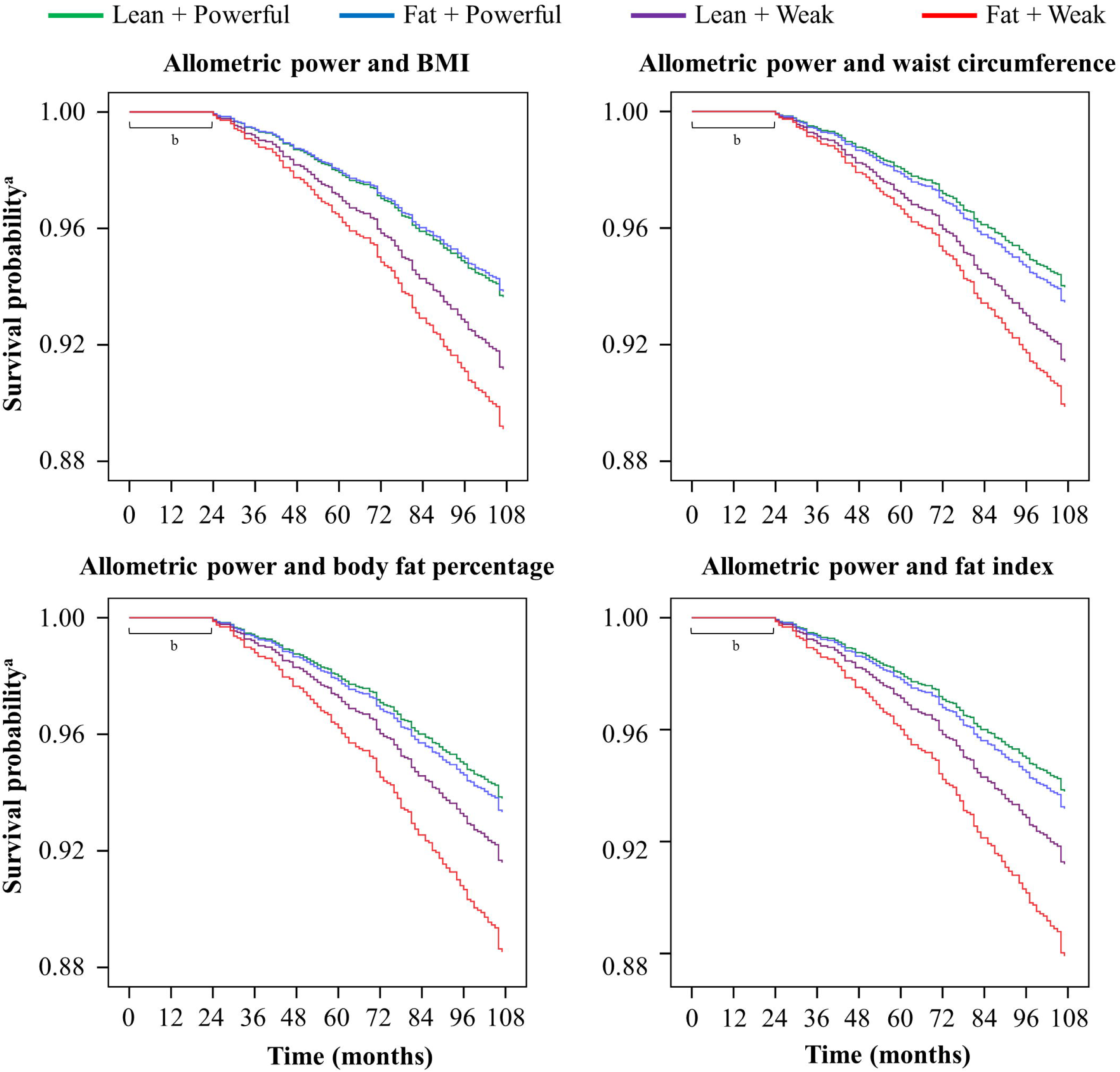
Survival probability for the different groups of adiposity and allometric power throughout the 9-year follow-up. ^a^Adjusted for age, sex, hypertension, smoking, and walking and sitting times. ^b^Deaths within the first 24 months were excluded to minimize bias from reverse causation.

Among the groups of allometric muscle power and BMI, hazard ratios of all-cause mortality were significantly reduced in the F+P (HR [95% CI] = 0.55 [0.31‒0.98]; p=0.044) and L+P (HR [95% CI] = 0.57 [0.33‒0.98]; p=0.043) groups when compared to the F+W group. No effect was reported in the L+W participants (HR [95% CI] = 0.80 [0.45‒1.44]; p=0.462) (Figure 2, panel A).

**Figure 2.**
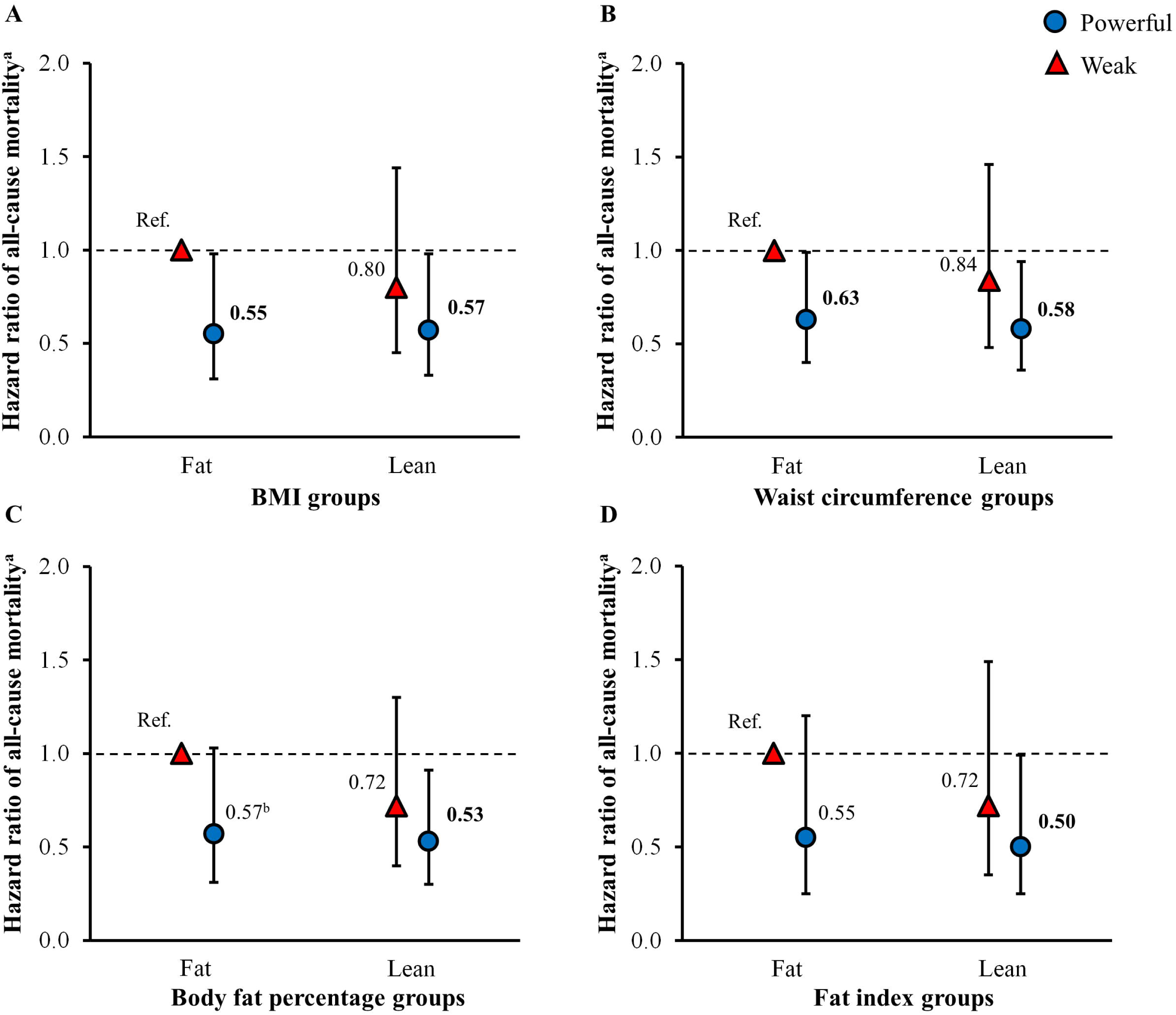
Hazard ratios with 95% confidence intervals of all-cause mortality among the different groups of allometric muscle power (powerful, *blue circles*, vs. weak, *red triangles*) and adiposity (fat vs. lean) according to BMI **(A)**, waist circumference **(B)**, body fat percentage **(C)** and fat index **(D)**. BMI, body mass index. Bold values denote statistically significant differences (*p*<0.05) compared to the reference category (fat and weak). ^a^Adjusted for age, sex, hypertension, smoking, and walking and sitting times. ^b^*p* = 0.060.

Similarly, when allometric power and waist circumference were regarded, all-cause mortality risk was reduced in F+P (HR [95% CI] = 0.63 [0.40‒0.99]; p=0.049) and L+P (HR [95% CI] = 0.58 [0.36‒0.94]; p=0.025) participants compared to the reference group, while no effect was found in the L+W group (HR [95% CI] = 0.84 [0.48‒1.46]; p=0.539) (Figure 2, panel B).

In terms of allometric power and body fat percentage, hazard ratios were again lower in the L+P group (HR [95% CI] = 0.53 [0.30‒0.91]; p=0.021), while a trend for a reduction was noted in the F+P participants (HR [95% CI] = 0.57 [0.31‒1.02]; p=0.060) in comparison to the F+W group. All-cause mortality risk was not significantly different in the L+W group (HR [95% CI] = 0.72 [0.40‒1.31]; p=0.721) when compared to the reference group (Figure 2, panel C).

Among the groups of allometric power and fat index, the L+P group was the only group showing a reduced all-cause mortality risk when compared to the F+W participants (HR [95% CI] = 0.50 [0.25‒0.99]; p=0.049), and thus, no effects were noted in the F+P (HR [95% CI] = 0.55 [0.25‒1.20]; p=0.133) and L+W (HR [95% CI] = 0.72 [0.35‒1.49]; p=0.370) groups (Figure 2, panel D).

### 3.3 Influence of relative muscle power on all-cause mortality

Finally, a reduced hazard ratio of all-cause mortality was noted in the participants with normal relative muscle power (HR [95% CI] = 0.60 [0.42‒0.86]; p=0.006) and high relative muscle power (HR [95% CI] = 0.52 [0.32‒0.86]; p=0.011) when compared with those with low relative muscle power (Figure 3).

**Figure 3.**
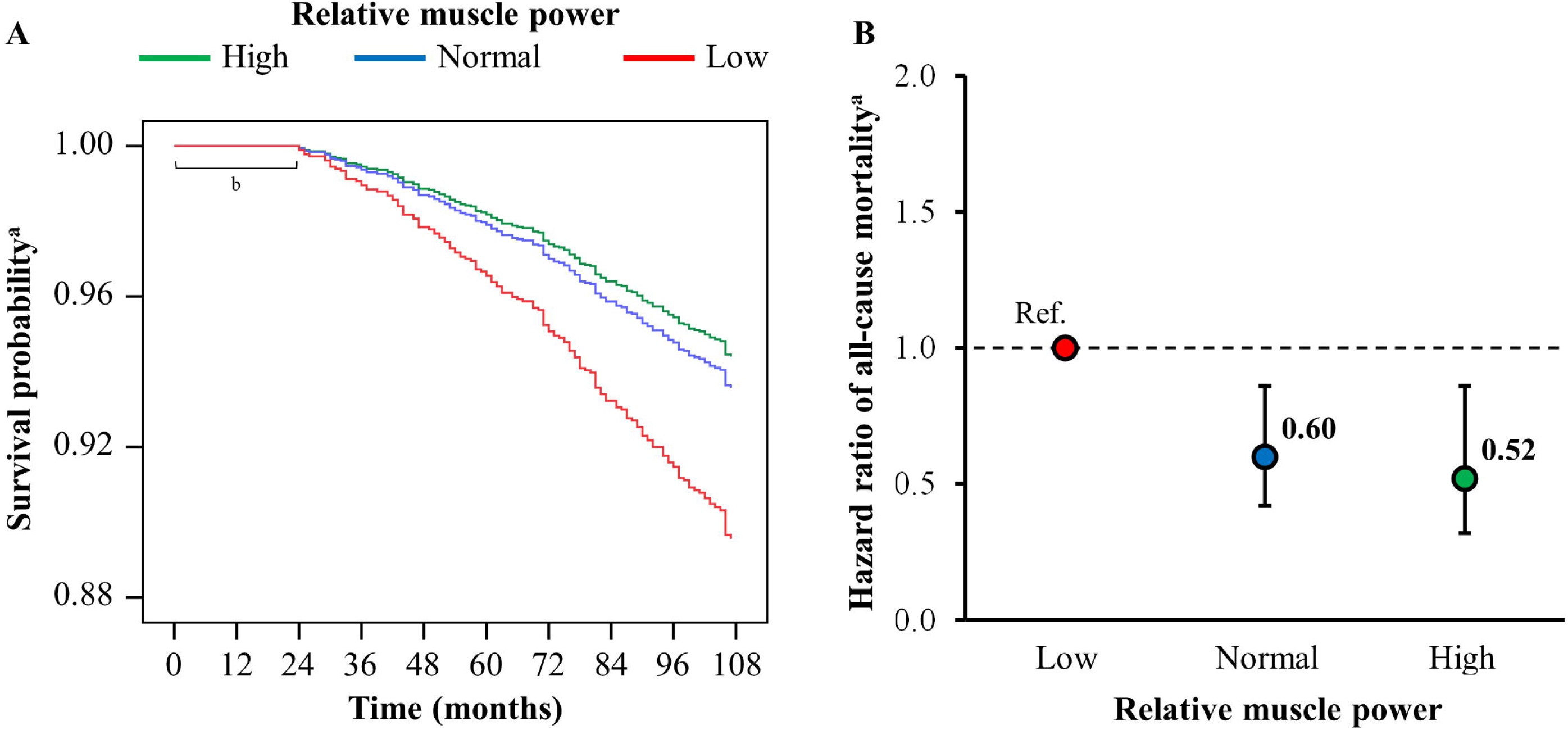
Survival probability **(A)** and hazard ratios with 95% confidence intervals of all-cause mortality **(B)** for the different groups of relative muscle power throughout the 9-year follow-up. Bold values denote statistically significant differences (*p*<0.05) compared to the reference category (low relative power). ^a^Adjusted for age, sex, hypertension, smoking, and walking and sitting times. ^b^Deaths within the first 24 months were excluded to minimize bias from reverse causation.

## 4. Discussion

The main findings of the present investigation were that the lean and powerful older adults presented a diminished all-cause mortality risk, while the fat but powerful participants showed a reduced all-cause mortality risk only when BMI and waist circumference were considered (all independently of age, sex, hypertension, smoking status and walking and sitting times). Moreover, normal and high levels of relative muscle power, which require appropriate combinations of allometric power and BMI, independently decreased all-cause mortality in older people.

### 4.1 Muscle power and all-cause mortality

Traditionally, cardiorespiratory fitness has been considered the cornerstone of physical fitness ^3^. In recent decades, muscle strength and resistance training have gained increased attention as numerous studies have reported their overall benefits ^18,19^, especially in older populations ^20-22^. Notably, it was found out that muscle power is a stronger predictor of functional ability in older people than muscle strength, muscle mass and aerobic capacity ^6, 23,24^. In addition, Metter et al. ^7^ demonstrated a significant association between low upper-limb absolute muscle power and mortality in middle-aged men after a 25-y follow-up, independently of muscle mass, strength and physical activity. However, due to i) absolute muscle power is positively correlated with height and ii) height progressively decreases across generations, the association between absolute muscle power and mortality may be – at least – partially explained by this aspect. Thus, we assessed allometric power (absolute power normalized to height squared) and confirmed the independent association of low muscle power with all-cause mortality in older men and women.

### 4.2 The ‘fat but fit/powerful’ paradox

Of note, in the current study being powerful seemed to be an independent requisite to live longer, while being lean was not. Previous studies have reported that body composition and physical fitness do not have equivalent influence on all-cause mortality risk. Ortega et al. ^3^ reported that being fit according to cardiorespiratory capacity decreased mortality risk in both men and women independently of the level of BMI, body fat percentage or waist circumference, while unfit individuals presented an increased mortality risk whether they were obese or not. The superiority of physical fitness over body composition regarding all-cause mortality risk was especially evident among older adults. Sui et al. ^5^ found that all-cause mortality risk was strongly modulated by cardiorespiratory fitness, while being obese or non-obese had no apparent effect on either fit or unfit older individuals. Similarly, we noted that the lean participants only presented a significantly decreased mortality risk when they were powerful as well, while all-cause mortality risk was generally significantly reduced in the powerful participants whether they were obese or not. However, there was an exception for the latter, since the obese group according to fat index values did not benefit from being powerful. Of note, BMI and waist circumference are used as proxies of fat mass or obesity, while body fat percentage also depends on the amount of other tissues (e.g. body fat percentage can be reduced by increasing muscle mass). Therefore, fat index is the adiposity marker that best and most independently reflects the amount of fat mass that an individual has, which may explain the discrepancies observed among adiposity measures. The harmful effects of obesity have been directly related to an excessive accumulation of fat mass, leading to the infiltration of adipocytes within other tissues, increased systemic low-grade inflammation, loss of function and augmented mortality ^25-27^. Thus, an excessive accumulation of fat mass (i.e. high fat index) would counteract the benefits of having adequate power levels. Nonetheless, all-cause mortality was lower (albeit non-significant) in the fat but powerful than in the lean and weak participants. Furthermore, among the adiposity indexes, the highest survival benefit when compared to the fat and weak group was observed in lean and powerful individuals according to fat index.

### 4.3 Relationship between relative muscle power and all-cause mortality

On the other hand, we also assessed the influence of relative muscle power on all-cause mortality among older people. Actually, relative muscle power combines a measure of physical fitness (allometric power) and a measure of adiposity (BMI), and is a more functionally relevant outcome when compared to absolute or allometric muscle power ^28,29^. In this sense, we observed normal and high levels of relative muscle power to protect against 9-year all-cause mortality independently of age, sex, hypertension, smoking, physical activity and sedentary time. Particularly, the survival benefits of relative muscle power seemed to plateau, and the protection conferred by high relative muscle power was relatively similar to that observed in the individuals with normal levels. The latter suggests that the relationship between relative muscle power and mortality is curvilinear. These findings help explain why the lean and powerful individuals generally did not exhibit a greater survival benefit than the fat and powerful participants. Relative muscle power increased progressively through the fat and weak, lean but weak, fat but powerful, and lean and powerful groups (weighted mean±SD = 1.75±0.40, 2.16±0.46, 2.90±0.58 and 3.25±0.61 W·kg^-1^, respectively). Therefore, above a certain level of relative muscle power no further survival benefits were noted. This curvilinear relationship has also been reported for the relationship between relative muscle power and physical performance in older adults ^30,31^.

## Limitations

Among the limitations of the study, it should be noted that the present investigation focused on independently living and non-institutionalized older adults, so our results may not apply to other older populations. On the contrary, the representativeness of our sample makes our results applicable to the general population of non-institutionalized Spanish older adults. In addition, a comprehensive assessment and recording of medical conditions was not performed, which could have improved our cox regression models. However, we did consider the main medical conditions and risk factors that have been related to all-cause mortality (hypertension, smoking, physical inactivity and obesity) ^32^. Finally, the procedure used in the present study to assess muscle power augments the applicability of our results, given that the STS muscle power test ^8,9^ is a valid, feasible and rapid test that can be performed in almost any global context, as long as a chair and a stopwatch are available.

### 5. Conclusions

Powerful older people exhibited a reduced 9-year all-cause mortality whether they were obese or not (according to BMI, waist circumference or body fat percentage), while being lean was a survival factor only when it was accompanied by the powerful condition. However, when fat index was regarded, reduced all-cause mortality risk was only observed in lean and powerful older adults, and so the survival benefits derived from being powerful were counteracted by the obese condition. Therefore, the ‘fat but fit/powerful’ paradox should be revisited using fat index as the adiposity marker. Finally, low relative muscle power assessed with the STS muscle power test was an independent risk factor for 9-year all-cause mortality in a representative sample of non-institutionalized Spanish older people.

## Data Availability

The data that support the findings of this study are available from the corresponding author on reasonable request.

## Contributorship

All authors were involved in data collection, analysis, writing and revision of the manuscript, and approved the final version submitted.

## Competing interests

The authors declare no competing interests.

## Funding

This work was supported by the Universidad de Zaragoza (UZ 2008-BIO-01); Gobierno de Aragón (Grant DGAIIU/1/20 to D.N.); Ministerio de Sanidad, Servicios Sociales e Igualdad (147/2011); Ministerio de Trabajo y Asuntos Sociales Sociales-IMSERSO (104/07); Centro Universitario de la Defensa de Zaragoza (UZCUD2016-BIO-01); Ministerio de Economía, Industria y Competitividad (DEP2016-78309-R); and the Biomedical Research Networking Center on Frailty and Healthy Aging (CIBERFES) and FEDER funds from the European Union (Grant CB16/10/00477).

## Ethical approval

The study was approved by the Clinical Research Ethics Committee of Aragón (18/2008) and the Ethical Committee of the University Hospital Fundación Alcorcón (50/2016),

## Patient and public involvement

Patients and/or the public were not involved in the design, or conduct, or reporting, or dissemination plans of this research.

## What are the findings?

- Older men and women with adequate levels of muscle power showed a 9-year survival benefit compared to their counterparts with low levels of muscle power, regardless of body mass index, waist circumference and body fat percentage levels.
- Obesity according to fat index (i.e. body fat normalized to height squared) mitigated the survival benefits provided by adequate levels of muscle power in older adults.
- The ‘fat but fit/powerful’ paradox was confirmed when adiposity was assessed by BMI, waist circumference and body fat percentage, but not by fat index.
- Low relative muscle power (i.e. power normalized to body mass) was an independent predictor of 9-year all-cause mortality among older people.

## How might it impact on clinical practice in the future?

- If possible, the assessment of fat index should be preferred over other adiposity markers due to it is more appropriate for quantifying the amount of body fat and was the only obesity marker that mitigated the survival benefits of muscle power.
- Nevertheless, in the fatness and fitness binomial, the greatest importance should be granted to the physical fitness component in terms of preventing mortality among older people.
- The assessment of low relative muscle power should be strongly encouraged in daily clinical practice given the previous evidence showing its detrimental consequences on physical performance and the novel evidence demonstrating its harmful effect on all-cause mortality among older people.

## Notes

### Competing Interest Statement

The authors have declared no competing interest.

### Funding Statement

This work was supported by the Universidad de Zaragoza (UZ 2008-BIO-01); Gobierno de Aragon (Grant DGAIIU/1/20 to D.N.); Ministerio de Sanidad, Servicios Sociales e Igualdad (147/2011); Ministerio de Trabajo y Asuntos Sociales Sociales-IMSERSO (104/07); Centro Universitario de la Defensa de Zaragoza (UZCUD2016-BIO-01); Ministerio de Economia, Industria y Competitividad (DEP2016-78309-R); and the Biomedical Research Networking Center on Frailty and Healthy Aging (CIBERFES) and FEDER funds from the European Union (Grant CB16/10/00477).

### Author Declarations

The study was approved by the Clinical Research Ethics Committee of Aragon (18/2008) and the Ethical Committee of the University Hospital Fundacion Alcorcon (50/2016).

